# Deleterious Variants Contribute Minimal Excess Risk in Large-Scale Testing

**DOI:** 10.1101/2024.10.21.24315653

**Authors:** Yen-Tsung Huang, En-Yu Lai, Jia-Ying Su, Hsueh-Ju Lu, Yen-Lin Chen, Jer-Yuarn Wu, Chun-yu Wei, Ling-Hui Li, Cathy S.-J. Fann, Hsin-Chou Yang, Chien-Hsiun Chen, Hung-Hsin Chen, Yi-Min Liu, Ming-Fang Tsai, Erh-Chan Yeh, Chih-Kuang Cheng, Yen-Po Wang, Nai-Fang Chi, I-Cheng Lee, Harn-Shen Chen, Yun-Cheng Hsieh, Yi-Chu Liao, Shao-Jung Hsu, Shuo-Ming Ou, Kuan-Lin Lai, Chung-Chi Lin, Yi-Jen Chen, Chia-Ming Chang, Peng-Hui Wang, Yung-Hung Luo, Yun-Ting Chang, Chih-Chiang Chen, Yu-Cheng Hsieh, Yi-Ming Chen, Tzu-Hung Hsiao, Ching-Heng Lin, Yen-Ju Chen, I-Chieh Chen, Chien-Lin Mao, Shu-Jung Chang, Yen-Lin Chang, Yi-Ju Liao, Chih-Hung Lai, Wei-Ju Lee, Hsin Tung, Ting-Ting Yen, Hsin-Chien Yen, Shy-Shin Chang, Yu-Sheng Chang, Ting-I Lee, Shauh-Der Yeh, Mei-Yi Wu, Ming-Shun Wu, Lung Wen Tsai, Cai-mei Zheng, Yu-Mei Chien, Tsung-Hsien Lin, Yen-Hsu Chen, Cheng-Che E. Lan, Jeng-Hsien Yen, Wen-Chen Liang, Te-Fu Chan, Shyh-Shin Chiou, Shih-Chang Chuang, Shang-Jyh Hwang, Yi-Jung Lin, Yu-Chuang Huang, Wan-Ru Li, Tsai-Chuan Chen, Wei-Ting Huang, Kuan-Chih Chen, Shin-Yee Lim, Yi-Shiuan Shen, Chia-Chia Huang, Chien-Hung Chen, Ya-Chung Tian, Chia-Ling Chen, Yao-Fan Fang, Ji-Tseng Fang, Yi-Hao Yen, Wei-Chi Wu, Wen-Shih Huang, Chi-Chin Sun, Mei-Jyh Chen, Ching-Hung Lin, Tsung-Hua Yang, Pei-Lin Lee, Ming-Yang Wang, Tsen-Fang Tsai, Tung-Hung Su, Jyh-Ming Liou, Shun-Fa Yang, Chia-Chuan Hsieh, Chih-Chien Sung, Feng-Chih Kuo, Shih-Hua Lin, Dueng-Yuan Hueng, Chien-Jung Lin, Hueng-Yuan Shen, Chang-Hsun Hsieh, Shinn-Zong Lin, Tso-Fu Wang, Tsung-Jung Ho, Pei-Wei Shueng, Chen-Hsi Hsieh, Kuo-Shyang Jeng, Gwo-Chin Ma, Ting-Yu Chang, Han-Sun Chiang, Yi-Tien Lin, Kuo-Jang Kao, Chen-Fang Hung, I-Mo Fang, Po-Yueh Chen, Kochung Tsui, Pui-Yan Kwok, Wei-Jen Yao, Shiou-Sheng Chen, Ming Chen, Chih-Yang Huang, Da-Wei Wang, Chun-houh Chen

## Abstract

DNA sequencing of patients with rare disorders has been highly successful in identifying “causal variants” for numerous conditions. However, there are many reports of healthy individuals who harbor these deleterious variants, leading to the concept of incomplete penetrance and doubt about the utility of genetic testing in clinical practice and population screening. As the deleterious variants are rare, the penetrance of these variants in the population is largely unknown. We analyzed the genetic and clinical data from 486,956 participants of the Taiwan Precision Medicine Initiative (TPMI) to determine the risk difference between those with and without deleterious variants. In all, we analyzed 292 disease-relevant variants and their clinical outcomes to assess their association. We found that only 15 variants show a risk difference exceeding 5% between those with or without the variants. In essence, 87.3% of deleterious variants exhibit minimal risk differences, suggesting a limited impact on the individual and population levels. Our analysis revealed increasing trends with age in six cardiovascular and degenerative diseases and bell-shaped trends in two cancers. Additionally, we identified three clinical outcomes exhibiting a dose-response relationship with the number of deleterious variants. Our findings show that large-scale testing of deleterious variants found in the literature is not warranted, except for those exhibiting large disease risk differences.

## 1 Introduction

A major advance in clinical genetics has been the identification of “causal variants” in genes responsible for rare genetic disorders by DNA sequencing of the patients.

The success of clinical sequencing has led to the discovery of new genes and deleterious variants for numerous conditions. About 170,000 genetic variants have been determined to be pathogenic or likely pathogenic in ClinVar [1], a database of genetic variation. While these “deleterious variants” fulfill consensus criteria for the designation, there are many reports of healthy individuals with deleterious variants who have never developed the associated conditions, and this leads to the concept of incomplete penetrance [2, 3].

Incomplete penetrance of deleterious variants reduces the predictive power of genetic testing, raising questions about its utility in clinical practice and population screening. Even for well-documented disease-associated variants, the level of risk may be overestimated [4] and this leads to unnecessary procedures, undue anxiety, and healthcare costs. For example, early cancer screening in individuals with deleterious variants in *BRCA1* and *BRCA2* genes can lead to diagnoses of early, curable stage cancers. However, deleterious variants in *BRCA1/2* do not always result in the development of breast or ovarian cancers due to low penetrance [5]. It is therefore important to determine the penetrance and further risk difference of each deleterious variant such that only highly penetrant variants with large risk difference are used in making clinical decisions. With such information, the benefits of genetic testing can be maximized while the undesired impact can be minimized.

There have been many studies conducted to quantify the penetrance of deleterious variants [6, 7]. In a recent study, Forrest et al. investigated the population-based penetrance of clinical variants and found that the estimated penetrance of pathogenic or loss-of-function variants was generally low [8]. However, the studies to-date are on European cohorts, and a comprehensive study focused on penetrance within the Asian population remains conspicuously absent. In this study, we analyzed genetic and clinical data from 486,956 participants in the Taiwan Precision Medicine Initiative (TPMI) to determine the penetrance of 292 selected deleterious variants. Those variants had robust genotyping precision and were annotated with clinical phenotypes. We further determined the risk difference based on penetrance and prevalence to infer the influence of deleterious variants on a population level. Our study is the largest of its kind and our results point to the limited utility of population screening of most of the 292 deleterious variants in asymptomatic Han Chinese individuals.

## 2 Results

### 2.1 Overview

We summarized the penetrance and prevalence of the 292 selected deleterious variants, along with their associated genes, disease annotations, phenotype categories, and age dependency in Figure 1. Detailed information on the 292 selected variants can be found in Supplementary Table S1, and the curation criteria can be found in Supplementary Figure S1. Since we mainly consider the influence of the deleterious variants on a population level, we present the risk difference (RD) of penetrance and prevalence on the same scale. Figure 2 shows that only 5.2% of the deleterious variants have large RDs (i.e., RD > 5%). If we consider inheritance patterns, then only 2.5% of the dominant variants and 8.9% of the recessive variants have large RDs. For this report, autosomal dominant (AD), X-linked dominant (XLD), mitochondrial (MT), and X-linked recessive in males (XLR-male) conditions are categorized as part of the dominant pattern, since one mutant allele is sufficient to induce the outcome; autosomal recessive (AR) and XLR in females (XLR-female) are part of the recessive pattern, since mutations in both copies of the gene in question are required for the outcome. Supplementary Figure S2 depicts the RD density of variants for different inheritance patterns.

**Fig. 1:**
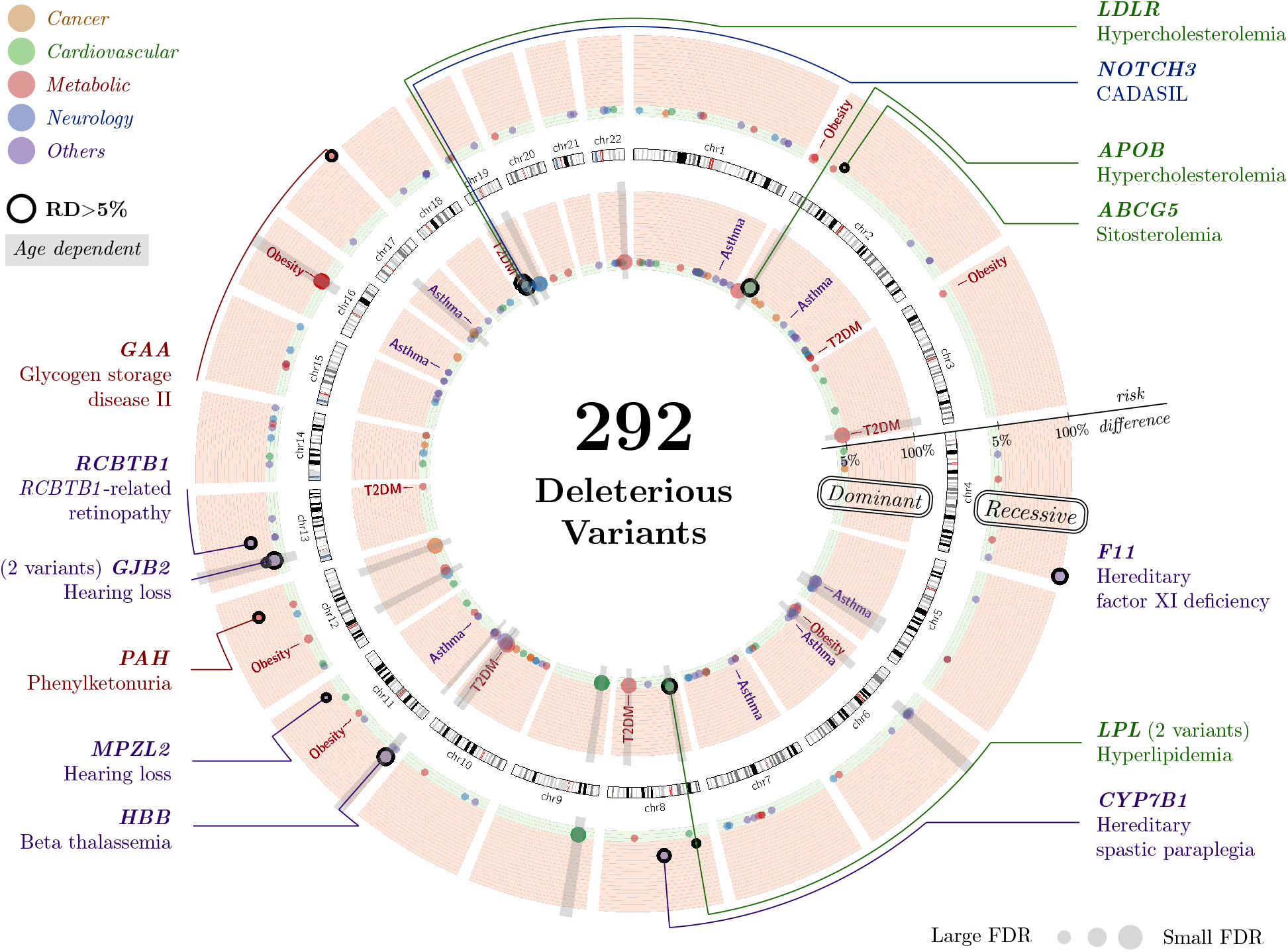
Overview of the 292 deleterious variants. We illustrated the characteristics of the 292 deleterious variants on the Circos plot. Among them, 163 variants of the dominant pattern were placed in the inner circle, and 123 variants of the recessive pattern in the outer circle. Notice that 54 variants are analyzed as both patterns and 60 recessive variants have no BB-carriers in our study. The circle’s height indicates the risk difference scale, where the < 5% region is colored green and > 5% colored red. The colors of the variants represent their phenotype categories; the age dependency and diseases with dose-response are also annotated. Gene names for the 15 variants with RD > 5% are listed.

**Fig. 2:**
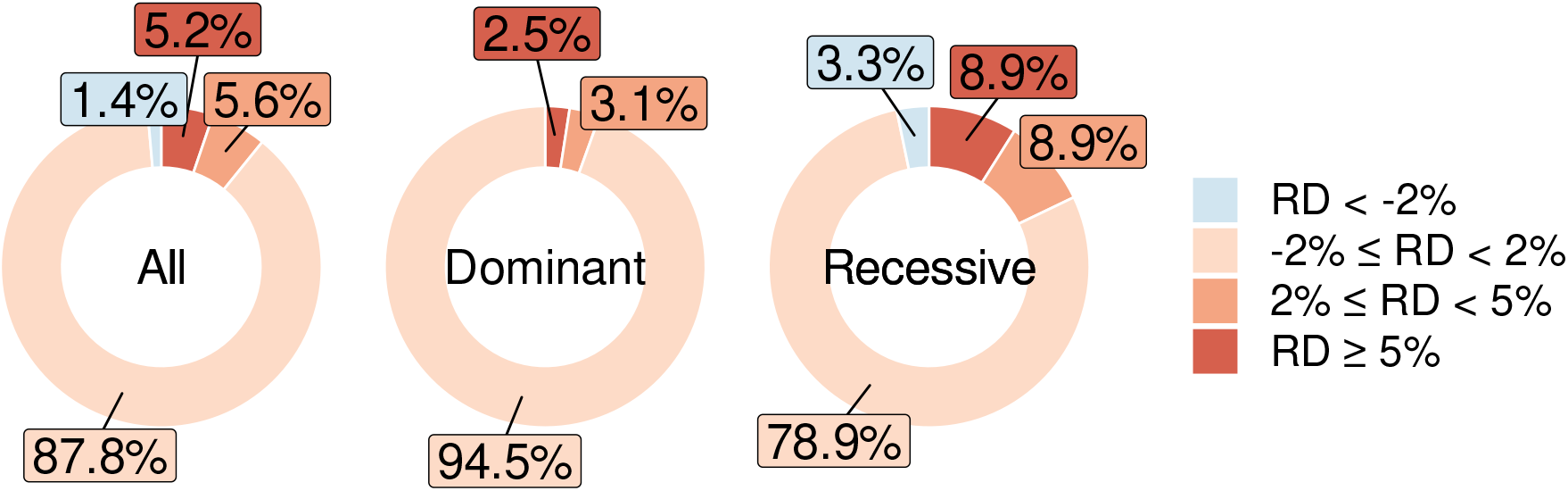
Donut charts of the deleterious variants. The inheritance patterns of 54 variants remain uncertain; in that case, we calculated these variants for both patterns (i.e., “as AR” or “as AD” in Supplementary Table S1). Then, 4 out of the 163 dominant variants (2.5%) had large RDs, and 11 out of the 123 recessive variants (8.9%) had large RDs.

Table 1 tabulates the exact numbers of affected individuals and their genotypes for the 15 gene variants where RD is > 5%. Of the 15 variants, 11 are AR variants: *F11* (rs770505620, RD=1.00), *GAA* (rs577915581, RD=1.00), *PAH* (rs76687508, RD=0.50), *RCBTB1* (rs777630688, RD=0.33), *CYP7B1* (rs200737038, RD=0.25), *MPZL2* (rs146689036, RD=0.21), *GJB2* (rs80338943, RD=0.19), *HBB* (rs34451549, RD=0.16), *LPL* (rs145657341, RD=0.14), *ABCG5* (rs119480069, RD=0.09), and *GJB2* (rs72474224, RD=0.07). Malfunction of these genes leads to relatively rare, and serious diseases, such as hereditary factor XI deficiency, Pompe disease, phenylketonuria, and *RCBTB1*-related retinopathy. Among the AR variants, 4 of them have a large false discovery rate (FDR), indicating the uncertainty of penetrance estimates due to the rare conditions. The 4 AD variants are in genes *LDLR* (rs730882109, RD=0.13), *APOB* (rs144467873, RD=0.12), *LPL* (rs371282890, RD=0.06), and *NOTCH3* (rs201118034, RD=0.06). Malfunction of these genes leads to relatively common, and mild diseases, such as hypercholesterolemia and hyperlipidemia. Of note, only 4 of the 15 deleterious variants are annotated in both InterVar and ClinVar, and 2 out of the 4 are also annotated in the TPMI expert panel. These results point to the diversity of clinical impact between different populations.

**Table 1:**
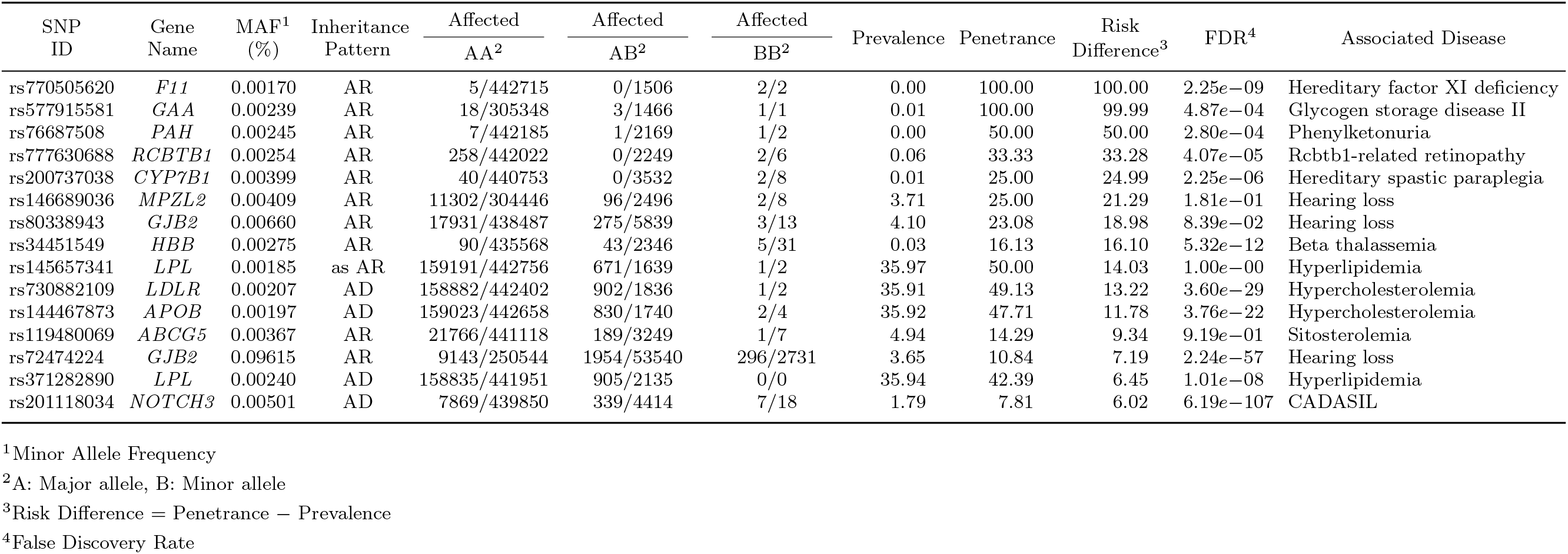
Deleterious variants with large risk differences. We listed the exact numbers to calculate the prevalence and penetrance for the 15 deleterious variants with RD > 5%. If the population scale is around 440000, the variant was identified from TPM1 and TPM2 chips. Otherwise, the variant was identified from the TPM2 chip alone. The nominal *p*-values were calculated by Fisher’s exact test (data not shown here, can be found in Supplementary Table S1), and the FDR values were calculated by BH method [9]. Variants with FDR>0.01 were highlighted.

We then investigated the 68 variants suggested by the TPMI expert panel (found in Supplementary Table 1). Considering the inheritance patterns of the 68 variants, 48 are AR variants and 6 are XLR-female variants (recessive pattern, 79.4% of total, mean RD=5.5%), while 8 are AD variants and 6 are XLR-male variants (dominant pattern, 20.6% of total, mean RD=2.9%). Only seven variants have large RD (5 of them are also found in ClinVar or InterVar). Variants rs730882109 in *LDLR* and rs144467873 in *APOB* are annotated only in the TPMI expert panel.

### 2.2 Age-stratified RD Patterns

As the results suggest, most variants have only small RDs between prevalence and penetrance, indicating that having the deleterious variant or not has only a limited influence on the outcome. Because some diseases may have a late age of onset, and our cohort has a wide range of ages, calculating RD without considering the effect of age may diminish the importance of certain variants. To investigate the influence of deleterious variants at different ages, we stratified the cohort into eight age groups for the analysis, and found three RD patterns to be influenced by age as illustrated in Figure 3.

**Fig. 3:**
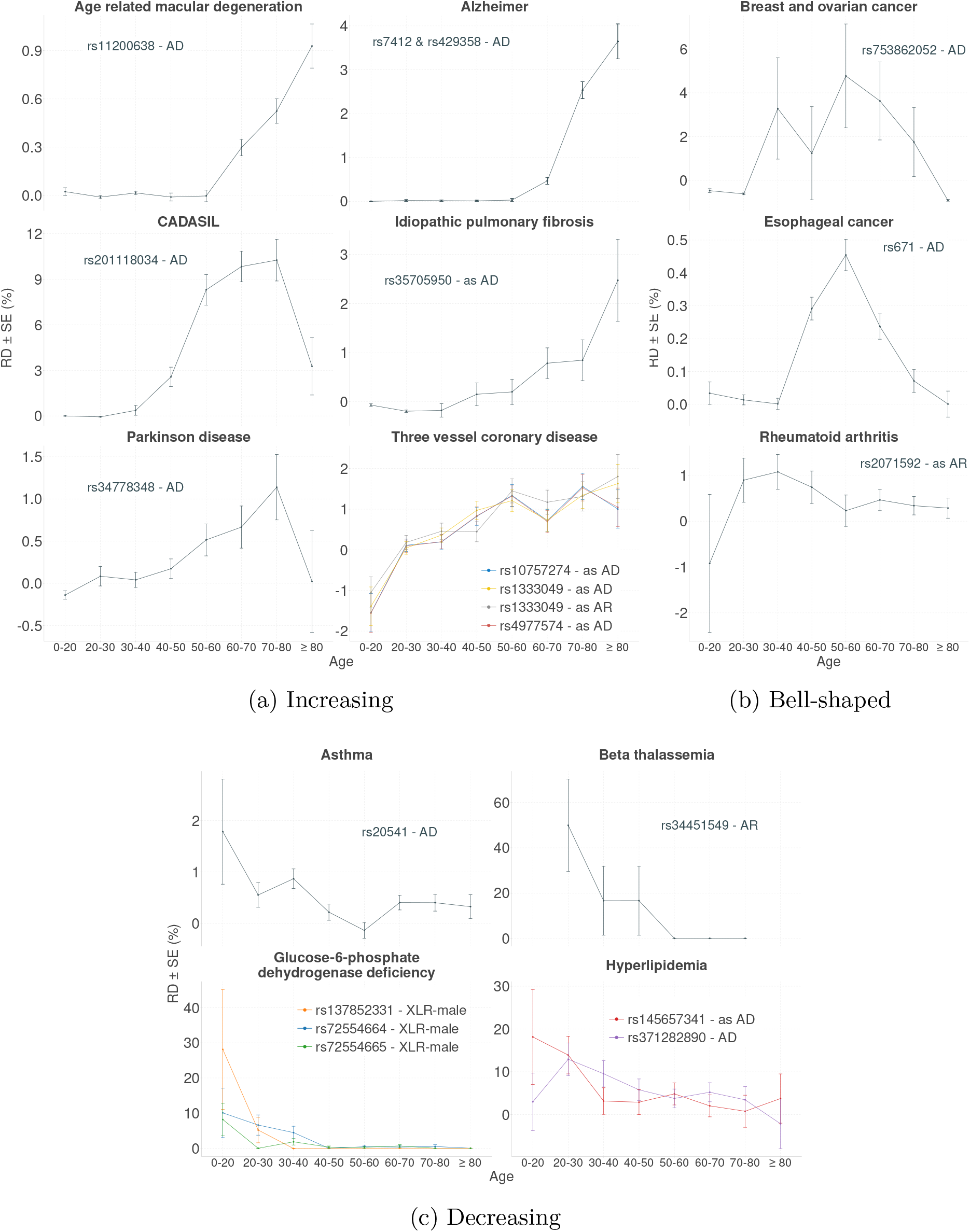
Three age-stratified RD patterns. We selected the variants with more than 30 carriers and analyzed the variants with RD > 5% or FDR < 0.01. For each variant, we calculated the *relative RD scale* among the age group (i.e., | max(RD) − min(RD)|*/*median(RD)).

Six clinical outcomes have an increasing RD pattern with age, indicating that the deleterious variants affect the older age groups. Although these variants have more influence on the elderly, their peak RD is still small, in general. Among the variants of increasing RD, only rs201118034 in *NOTCH3* reaches RD> 10% in CADASIL (Cerebral Autosomal Dominant Arteriopathy with Subcortical Infarcts and Leukoencephalopathy) at ages 60-80. Other variants with higher RD for the elderly have limited influence on disease (i.e., the highest RD in the onset group is still < 5%). Two cancers have a bell-shaped RD pattern, indicating that middle-aged individuals are more vulnerable to deleterious variants. We also found that individuals in their early adulthood are more vulnerable to the deleterious variant leading to rheumatoid arthritis. Among the three bell-shaped variants, only rs753862052 in *RAD51D* has a large RD for breast and ovarian cancer. Four clinical outcomes have a decreasing pattern for RD, indicating that younger people are more vulnerable to deleterious variants. Among the variants of decreasing RD, only rs34451549 in *HBB* is a recessive variant and has a large RD at ages 20-30; this variant is associated with beta thalassemia. The other six are dominant variants: rs371282890 and rs145657341 in *LPL* have large risk differences in hyperlipidemia at ages 0-30; rs137852331, rs72554664, and rs72554665 in *G6PD* also have large risk differences in glucose-6-phosphate dehydrogenase deficiency at ages 0-30. Figure 3 shows the outcomes with clear differences between the age groups (i.e. relative RD scale = | max(RD) − min(RD)|/median(RD) ≥ 3); Supplementary Figure S4 shows the RD patterns of all the outcomes.

### 2.3 Dose–Response Relationship between Variants and Risks

Clinical outcomes usually involve intricate mechanisms; therefore, disease syndromes are generally polygenic. To examine whether those with more deleterious variants are more likely to develop the disease, we investigated the dose-response relationship between the number of variants and the risk of disease. Figure 4 shows that the risks of developing obesity, asthma, and type 2 diabetes mellitus increase with the number of variants possessed. However, comparing with non-carrier, having more than 5 variants only increases 1-3% of risk. Additionally, obesity and asthma have higher risks in the younger population, but type 2 diabetes mellitus has a higher risk in the older population. Type 2 diabetes mellitus is also the only disease that has a conspicuous risk difference between the younger and the older population. Other diseases without a clear dose-response relationship can be found in Supplementary Figure S5.

**Fig. 4:**
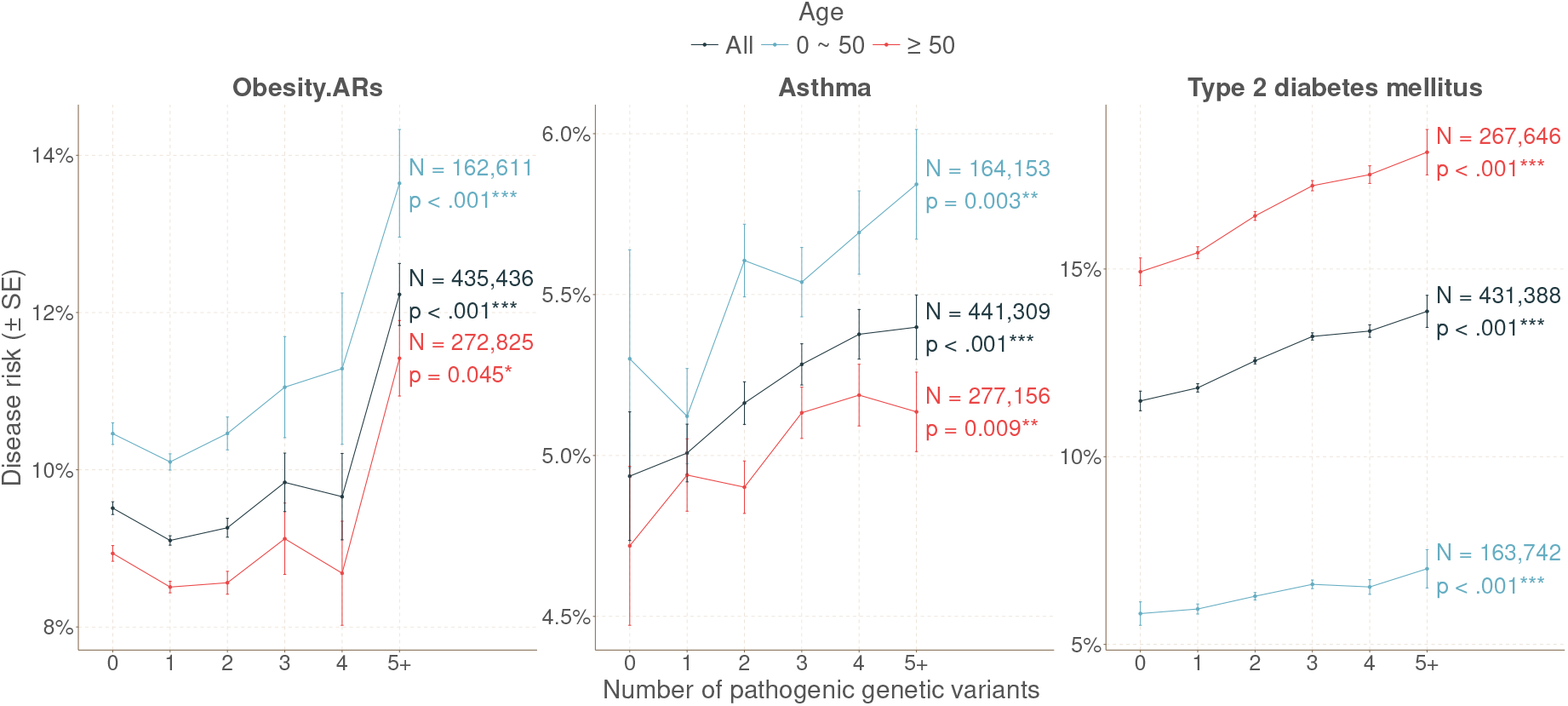
Dose–Response relationship between Variants and Risks. Three diseases associated with > 5 deleterious variants with significant *p*-values of dose-response relationship. The *p*-values are calculated using the Cochran-Armitage trend test. Compared to non-carriers (0), the risk of developing the disease increases by 1-3% for those possessing more than 5 variants (5+).

### 2.4 Phenotype Category

Supplementary Figure S3 shows the violin plots of deleterious variants on penetrance and prevalence for five phenotype categories. Among the 15 variants in Table 1, 5 are related to cardiovascular diseases (belong to *LDLR, APOB, ABCG5*, and two variants in *LPL*); two to metabolic diseases (*GAA* and *PAH*); and one to neurology diseases (*NOTCH3*). Among the 55 variants with FDR < 0.01 in Supplementary Table S1, 2 are related to cancer, 10 to cardiovascular diseases; 26 to metabolic diseases; and 4 to neurology diseases. The two variants related to cancers, *RAD51D* and *ALDH2*, are the variants with bell-shaped age dependency in Figure 3; and the four variants related to neurology diseases, *NOTCH3, LRRK2*, and two variants in *APOE*, all show an increasing age effect. Variants related to the well-known cancer predisposition gene *BRCA1* and *BRCA2* are not included in our initial 292 variants because they do not meet the MAF and NGS concordance criteria (Supplementary Figure S1).

## 3 Methods

### 3.1 Study population

The TPMI is a nationwide research project encompassing 486,956 individuals with both genetic profiles and clinical data from 16 hospital systems across Taiwan [10–12]. Participants provided their informed consent for SNP genotyping and access to their electronic medical records (EMRs) at the recruitment hospitals. The dataset integrates comprehensive demographic, medical, and genotyping information. The TPMI participants were genotyped on the customized Taiwan Precision Medicine chips, TPM1 and TPM2 Axiom Arrays (Thermo Fisher Scientific), designed jointly with the Taiwan Biobank. Whole-genome genotyping was performed using the TPM1 array for 165,666 participants and the TPM2 array for 321,360 participants, with 70 individuals having data available from both arrays. The TPM1 and TPM2 chips target 686,439 and 725,936 genetic loci, respectively, designed specifically to capture the genetic diversity of the Han Chinese population. This study was granted by the Ethics Committees of Academia Sinica and the participating hospitals of the TPMI. Rigorous quality control measures were implemented to exclude samples with discrepancies between genetic and documented sex, as well as any duplicates.

### 3.2 Variant curation and annotation

To study the clinical impact and penetrance of potential actionable genomic findings of the overlapping markers on TPM1 and TPM2 Axiom Array, we curated the clinically actionable variants, including pathogenic (P), likely pathogenic (LP) variants and risk variants annotated by ClinVar and InterVar, and known disease-related variants in Taiwanese chosen by a TPMI expert panel. The P/LP variants were extracted from ClinVar [1] (2023-06-17 release) and InterVar [13] (2022-06-13 release), and the risk variants annotated by ClinVar were also included. The list of the known disease-related variants in Taiwanese was curated by TPMI experts, including variants used for the molecular diagnosis of diseases, founder mutations of common inheriting diseases, and common disease-relevant variants.

The genes of all the variants studied were mapped onto ref gene from the UCSC genome browser [14] (https://genome.ucsc.edu/), and the related disease were obtained from the American College of Medical Genetics Secondary Findings v3.2 list [15] (ACMG), ClinGen Actionability Reports [16], ClinVar, Online Mendelian Inheritance in Man [17] (OMIM, https://www.omim.org/) and Orphanet [18] (https://www.orpha.net/consor/cgi-bin/index.php). The inheritance patterns of the diseases were extracted from OMIM, Orphanet, and MedlinePlus [19] (https://medlineplus.gov/). The Phenotype Categories of the diseases were obtained from ACMG, Orphanet, and MedlinePlus. The ICD-10 codes of each pathological condition were assigned based on the information from Health Promotion Administrative (HPA), Taiwan Foundation of Rare Disorders (TFRD), The American Academy of Ophthalmology (AAO), Orphanet, and TPMI clinical experts. The compilation includes only those variants that align with both the disease names and ICD-10 codes, accompanied by a documented mode of inheritance. The 292 variants in the final list (Supplementary Table S1) required an overall call rate > 95%, minor allele frequency (MAF) ≥ 0.1% (the genotyping accuracy cannot be guaranteed at lower MAF), and next generation sequencing (NGS) concordance ≥ 0.8, and selection criteria are shown in Supplementary Figure S1.

### 3.3 Phenotyping of patients

To pinpoint patients diagnosed with specific diseases, we employed ICD-10 codes. The case group is defined based on the inclusion of a minimum of one diagnostic code entry within the medical documentation, originating from outpatient, inpatient, emergency, surgical, or medical imaging report records. Individuals who do not meet the specified criteria for any of the phenotypes are classified as the control group. For five specific phenotypes—hyperlipidemia, hypercholesterolemia, obesity, type 2 diabetes mellitus, and CADASIL—a mere presence of diagnostic codes was considered insufficient. Instead, these conditions required further qualifications involving additional test results or specific findings in imaging reports to be classified as cases.

The case definition for hyperlipidemia and hypercholesterolemia is not only based on their respective ICD-10 codes but also required at least one instance of documented biochemical markers showing total cholesterol (TC) ≥ 200 mg/dl, triglycerides (TG) ≥ 130 mg/dl, or low-density lipoprotein cholesterol (LDL-C) ≥ 190 mg/dl. Similarly, for obesity, a body mass index (BMI) of ≥ 30 kg/m^2^ is required in addition to the diagnostic codes. Type 2 diabetes mellitus cases are confirmed with both the ICD-10 codes and a hemoglobin A1c (HBA1c) test result > 6.5%. CADASIL requires not just the ICD-10 codes but also the presence in imaging reports of terms such as ”lacune”, ”lacunar infarction”, ”small vessel”, ”white matter hyperintensity”, ”leukoaraiosis”, ”microbleed”, ”small infarct”, or ”white matter lesion” documented at least once in brain computed tomography (CT) or brain magnetic resonance imaging (MRI) scans.

### 3.4 Statistical analysis

#### Definitions of Carrier and Non-Carrier, and Quantification of Penetrance and Prevalence

The estimation of penetrance focused on quantifying disease prevalence among carriers, while prevalence indicated the proportion of disease occurrence among non-carriers. The definition of “carrier” and “non-carrier” varied depending on the specific mode of inheritance for each disease [8]. In the context of inheritance modes such as autosomal dominant (AD), non-specific patterns (either dominant or recessive) X-linked (XL), X-linked dominant (XLD), and mitochondrial (MT), individuals carrying at least one minor allele were designated as “carriers”, while the remaining individuals were referred to as “non-carriers”. For autosomal recessive (AR), individuals required two minor alleles to be considered “carriers”. Concerning X-linked recessive (XLR) inheritance, we performed gender-specific analyses. For males, carriers of XLR were defined as individuals with a deleterious allele, while for females, carriers of XLR were those with homozygous deleterious alleles; the rest were classified as non-carriers.

### Assessing the Impact of Genetic Variants on Phenotypes

To ascertain the significance of variant effects on their respective diseases, we employed Fisher’s exact test using a 2 *×* 2 contingency table (comprising carriers/non-carriers versus affected/unaffected individuals). For a deeper insight into the variant effect, we employed the risk difference (the difference between penetrance and prevalence) to investigate the significance of each variant. All graphical representations and statistical analyses were performed using R version 4.3.3.

### Donut charts of RD distribution

To elucidate the distribution of RDs across the studied genetic variants, we utilized donut charts. These charts categorize the variants into defined RD ranges and visually represent these categories as proportions of the total.

Variants were categorized into four groups based on their RD values:

- **RD** < **-2%:** Variants in this group suggest a decreased prevalence of disease among carriers compared to non-carriers.
- **-2%** ≤ **RD** < **2%:** This range includes variants with negligible impact on disease risk.
- **2%** ≤ **RD** < **5%:** Variants in this category have a moderate increase in disease prevalence among carriers.
- **RD** ≥ **5%:** These variants are associated with a substantial increase in disease risk, highlighting their potential importance in genetic screening and clinical interventions.

The analysis covered 292 unique variants, of which 232 displayed distinct RD values. Among these, the inheritance patterns were classified based on the variant’s impact in monoallelic (dominant) or biallelic (recessive) conditions:

- **Dominant (163 variants):** Includes variants associated with autosomal dominant (AD), mitochondrial (MT), X-linked (XL), and X-linked recessive in males (XLR-male) inheritance patterns. For these variants, possessing one mutant allele is enough to influence the phenotype.
- **Recessive (123 variants):** Comprises variants associated with autosomal recessive (AR) and X-linked recessive in females (XLR-female) inheritance patterns. These variants require two mutant alleles to affect the phenotype.

### Age-stratified RD analysis

Understanding the age-specific impacts of genetic variants on their associated phenotypes is crucial, given that many diseases have age-dependent onsets and progressions. By conducting age-stratified RD analyses, we aim to reveal whether certain variants have disproportionate effects on disease risk at particular stages of life. This is particularly pertinent for conditions with known age of onset, where variants might exhibit significant influence in specific age brackets. First, we selected variants from the cohort that exceeded 30 carriers, targeting those with an RD greater than 5% or an FDR less than 0.01. This stringent selection process was employed to isolate variants that are statistically significant or carry potential clinical importance. Inclusion of variants with an FDR less than 0.01 allows for the consideration of those that are statistically significant but might not show a broad RD greater than 5%. This inclusion criteria ensures we do not overlook variants that, while not broadly impactful, may exhibit significant RD variations in restricted age groups. For each selected variant, we computed the relative RD scale among the age groups, defining it as | max(RD)−min(RD)|*/*median(RD). This measure helped us to identify and illustrate the extent of RD variation across different age categories. Diseases with a relative RD scale larger than 3 were emphasized in Figure 3, signifying considerable fluctuations in risk which may provide insights into age-specific genetic influences. By adopting a threshold that brings out the most pronounced RD variations, we aimed to prioritize findings that could indicate important age-related changes in disease risk profiles. Comprehensive results, capturing the entire spectrum of relative RD scale values, have been detailed in Supplementary Figure S4 for extended analysis and complete representation of the data.

#### Dose–response analysis between variants and risk

We conducted dose–response analyses to investigate the potential correlation between the number of genetic variants and the disease risk. This analysis included diseases associated with more than five variants identified by both TPM1 and TPM2 chips. The statistical significance of the trends was assessed using the Cochran-Armitage trend test, which is specifically designed for ordered alternatives, making it suitable for evaluating trends across ordered groups of variant counts. The primary goal was to determine whether an increase in variant count is associated with an increase in disease risk, particularly examining if this trend persists across different age groups. Given the onset of many genetic disorders varies significantly with age, it is crucial to understand how these associations change with age. Therefore, we analyzed both overall trends and stratified the data by age groups (above and below 50 years) to assess whether the observed dose–response relationships were consistent across different age groups. The diseases associated with more than five variants were selected for detailed analysis, including obesity (analyzed as ADs and ARs), breast and ovarian cancer (analyzed as ADs and ARs), asthma, Type 2 diabetes mellitus, Glucose-6-phosphate dehydrogenase deficiency, and retinitis pigmentosa (analyzed as ADs and ARs). The terms as ADs and as ARs refer to analyses conducted under the assumption that the unclear inheritance patterns of some variants were either autosomal dominant (AD) or autosomal recessive (AR), respectively. Diseases such as Glucose-6-phosphate dehydrogenase and retinitis pigmentosa were included in the initial analysis phase but were excluded due to the absence of patients with more than one variant. Only diseases that demonstrated a significant overall dose–response relationship were highlighted in Figure 4, ensuring the findings reflect substantial and clinically relevant variant–disease associations. All analysis results, including both significant and non-significant outcomes, are detailed in supplementary Figure S5.

## 4 Discussion

With the ever-increasing number of deleterious variants identified by clinical sequencing of patients with rare diseases, and the push to sequence “everyone” to incorporate genetic information in clinical practice, assessing the real disease risk in asymptomatic individuals with deleterious variants is of great clinical importance. As the deleterious variants are usually rare, only large cohort studies have sufficient power to provide evidence for disease risk levels relevant in clinical practice. In this study, we compared the penetrance and prevalence of 292 deleterious variants in a cohort of 486,956 individuals in the TPMI and found that only 15 variants have RD > 5% between those with and without the deleterious variants. Specifically, 94.8% of the deleterious or loss of function variants found in the databases have limited impact in the Han Chinese population. Even if we only consider the 68 clinical variants suggested by the TPMI expert panel, 89.7% of the variants have RD < 5%. Similar results were presented in a previous study using BioMe and UK Biobank, where 89% of the variants have RD < 5% [8]. These results clearly indicate that most deleterious variants found in patients with rare diseases (and published in peer reviewed journals or submitted to databases such as ClinVar) do not exhibit a significant effect on asymptomatic individuals or the population as a whole.

For the small number of deleterious variants with significant risk differences, most of them cause severe AR diseases. For example, the top three AR variants are related to hereditary factor XI deficiency, Pompe disease, and phenylketonuria, all of which lack curative treatments and rely on symptomatic therapies to prevent excessive bleeding [20], glycogen buildup [21], or irreversible mental disability [22]. Additionally, the average risk difference is notably higher in recessive diseases (RD = 0.0327) compared to dominant diseases (RD = 0.0040), highlighting the considerable influence of inheritance patterns on the transmission of deleterious variants in the population. Despite the severity of AR-associated diseases, affected individuals represent a small fraction of the population, as the recessive pattern requires two alleles for trait manifestation. For example, only two out of approximately four hundred thousand participants in the TPMI cohort inherit the top-ranked homozygous variant rs770505620. On the contrary, three of the four autosomal dominant (AD) variants in Table 1 are associated with relatively mild conditions such as hypercholesterolemia and hyperlipidemia compared to AR-inherited diseases.

The variant rs201118034 (p.R544C) (Table 1) deserves our attention for several reasons. First, as an AD variant in the *NOTCH3* gene, an individual who inherited one copy of the mutated gene from either parent is supposed to develop the associated adult-onset condition, CADASIL, a hereditary small vessel disease of the brain with stroke as a defining symptom [23, 24]. Second, approximately 1% of individuals in Taiwan carry the rs201118034 variant (1% in the TPMI cohort, 0.9% in the Taiwan Biobank cohort [23]), and now our result showed that carriers of rs201118034 variant face a risk difference of roughly 6% compared to non-carriers (Table 1), the risk difference even reaches to 10% at ages 60-80 (Figure 3(a)). Because there is no cure for CADASIL, genetic counseling and health management measures that delay disease onset are the best course of action to reduce the burden of disease in the population. Third, mutations in the *NOTCH3* gene can also lead to cognitive impairment and other neurological deficits beyond stroke, underscoring the multifaceted impact of this genetic variant on cognitive functions [25, 26]. Fourth, understanding and monitoring the implications of rs201118034 is essential for effective management and intervention strategies to mitigate the risks associated with CADASIL and its broader neurological manifestations.

In addition to CADASIL, Alzheimer’s and Parkinson’s disease also showed conspicuous increases in risk differences with age (Figure 3(a)), suggesting that not only aging itself plays a key risk factor for neurodegenerative diseases [27, 28], but the risk increases even more with age if one carries certain deleterious variants. We can draw a similar conclusion on three vessel coronary disease, as the risk differences of four related variants also increase with age (Figure 3(a)). A similar study of a multicenter case cohort also observed an increase in penetrance with age in cardiovascular disease [29]. The rationale behind the increasing RD pattern can be easily comprehended. In the context of neurodegenerative diseases, the accumulation of misfolded proteins, oxidative stress, and inflammation are prominent features of aging brains [27, 28]. Similarly, in cardiovascular diseases, the arteries become less flexible and more prone to plaque build-up with age, leading to reduced blood flow and an increased risk of cardiovascular events [30]. Our results pointed out that carrying certain deleterious variants worsens the clinical conditions while aging. Cancers, in general, also have increased risks with age [31], but Figure 3(b) depicted a bell-shaped pattern for breast and ovarian cancer, and esophageal cancer. We noticed that screening practices may also influence the observed risk across age groups and may result in bell-shaped or decreasing RD patterns. For example, in Taiwan, the newborn screening for *G6PD* deficiency became a regular test in 1987, and the screening rate reached 99% in 1996; breast cancer is also routinely screened since the government provides free mammography screening for women at 45-70 ages [32].

Finally, we explored the dose-response relationship between the number of variants and disease risk. While previous studies primarily focused on this relationship within different alleles of the same gene, comparing the effects of heterozygous and homozygous variants, comprehensive analyses encompassing both genome-wide and phenome-wide perspectives have been limited. In our study, we identified three polygenic diseases — obesity, asthma, and type 2 diabetes mellitus — that exhibit a dose-response relationship with the number of associated variants. Among the 18 variants associated with obesity, the top 1 to 12 belongs to the *FTO* gene, which not only increases body and fat mass but also affects food intake [33]. While none of these 18 variants individually demonstrate large risk differences, individuals carrying 5 or more variants exhibit a significantly higher average risk difference compared to non-carriers, consistent with previous research findings [34]. Similar observations were made in asthma, where 9 variants, including 3 belonging to interleukin genes, were associated with the disease, supporting the presence of a polygenic effect, as suggested in previous studies [35]. Type 2 diabetes mellitus also displays a polygenic nature with a dose-response effect, where different sets of variants can lead to distinct etiologies [36]. Among the 9 associated variants, the top variants belong to *TCF7L2, IGF2BP2*, and *SLC30A8*, genes that influence insulin secretion [37–39]. Our findings further reveal that the elderly population exhibits a stronger dose-response effect on Type 2 diabetes than younger individuals, highlighting the intricate relationship between aging and the penetrance of variants.

## 5 Conclusion

With the exception of 15 deleterious variants, our analysis of 292 deleterious variants in 486,956 Han Chinese individuals in Taiwan shows that the vast majority of deleterious variants identified in patients with rare diseases confer minimal added risk to those with the risk variant in the general population. This finding has several implications. First, genetic testing of most deleterious variants found in ClinVar or the literature in the general population will have no impact on individual or population health management. Second, identifying protective or modifying genetic factors in asymptomatic individuals with deleterious variants will likely lead to useful insights into the diseases in question. Third, in several adult-onset diseases, the risk difference exhibits an age-related increasing trend. Fourth, three conditions display a dose-response relationship with the number of deleterious variants present in a patient. Overall, we have conducted the largest study on the penetrance of deleterious variants to-date and laid the foundation for risk assessment for these variants in the clinical setting.

## Data Availability

The genotyping and electronic medical record (EMR) data analyzed in this study are from the Taiwan Precision Medicine Initiative (TPMI) with proper approval from the TPMI Data Access Committee. In compliance with the confidentiality laws governing genetic and health data in Taiwan, the de-identified TPMI data are kept in a secure server at the Academia Sinica and not released to the public. Researchers requesting access to the individual genotyping and EMR data can do so on a collaborative basis. Instructions on requesting access to the data can be found on the TPMI official website (https://tpmi.ibms.sinica.edu.tw/www/en/).

## Supplementary information

Supplementary information includes a list of 292 deleterious variants in a csv file (Table S1), and a pdf file containing demographic characteristics of the TPMI Biobanks (Table S2), variant curation of the 292 deleterious variants (Figure S1), RD distribution of the 292 deleterious variants (Figure S2), Violin plots of the 292 deleterious variants (Figure S3), age-stratified RD patterns (Figure S4), and dose-response relationship of the selected variants and disease risks (Figure S5).

## Declarations

### Ethics approval

This study was approved by the Institutional Review Boards of 17 institutes: 1. Academia Sinica (AS-IRB01-18079), 2. Taipei Veterans General Hospital (2020-08-014A), 3. National Taiwan University Hospital (201912110RINC), 4. Tri-Service General Hospital (2-108-05-038), 5. Chang Gung Memorial Hospital (201901731A3), 6. Taipei Medical University Healthcare System (N202001037), 7. Chung Shan Medical University Hospital (CS19035), 8. Taichung Veterans General Hospital (SF19153A), 9. Changhua Christian Hospital (190713), 10. Kaohsiung Medical University Chung-Ho Memorial Hospital (KMUHIRB-SV(II)-20190059), 11. Hualien Tzu Chi Hospital (IRB108-123-A), 12. Far Eastern Memorial Hospital (110073-F), 13. Ditmanson Medical Foundation Chia-Yi Christian Hospital (IRB2021128), 14. Taipei City Hospital (TCHIRB-10912016), 15. Koo Foundation Sun Yat-Sen Cancer Center (20190823A), 16. Cathay General Hospital (CGH-P110041), 17. Fu Jen Catholic University Hospital (FJUH109001).

### Data availability

The genotyping and electronic medical record (EMR) data analyzed in this study are from the Taiwan Precision Medicine Initiative (TPMI) with proper approval from the TPMI Data Access Committee. In compliance with the confidentiality laws governing genetic and health data in Taiwan, the de-identified TPMI data are kept in a secure server at the Academia Sinica and not released to the public. Researchers requesting access to the individual genotyping and EMR data can do so on a collaborative basis. Instructions on requesting access to the data can be found on the TPMI’s official website (https://tpmi.ibms.sinica.edu.tw/www/en/).

### Author contribution

#### Conceptualization

Yen-Tsung Huang, Pui-Yan Kwok, En-Yu Lai, Jia-Ying Su, Chun-yu Wei

#### Formal analysis

Jia-Ying Su, En-Yu Lai, Yen-Tsung Huang, Chun-yu Wei

#### Investigation

Hsueh-Ju Lu, Yen-Lin Chen, Chih-Kuang Cheng, Kuan-Chih Chen, Shin-Yee Lim, Yi-Shiuan Shen, Chia-Chia Huang, Yen-Po Wang, Nai-Fang Chi, I-Cheng Lee, Harn-Shen Chen, Yun-Cheng Hsieh, Yi-Chu Liao, Shao-Jung Hsu, Shuo-Ming Ou, Kuan-Lin Lai, Chung-Chi Lin, Yi-Jen Chen, Chia-Ming Chang, Peng-Hui Wang, Yung-Hung Luo, Yun-Ting Chang, Chih-Chiang Chen, Yu-Cheng Hsieh, Yi-MingChen, Tzu-HungHsiao, Ching-Heng Lin, Yen-Ju Chen, I-Chieh Chen, Chien-Lin Mao, Shu-Jung Chang, Yen-Lin Chang, Yi-Ju Liao, Chih-Hung Lai, Wei-Ju Lee, Hsin Tung, Ting-Ting Yen, Hsin-Chien Yen, Shy-Shin Chang, Yu-Sheng Chang, Ting-I Lee, Shauh-Der Yeh, Mei-Yi Wu, Ming-Shun Wu, Lung Wen Tsai, Cai-mei Zheng, Yu-Mei Chien, Tsung-Hsien Lin, Yen-Hsu Chen, Cheng-Che E. Lan, Jeng-Hsien Yen, Wen-Chen Liang, Te-Fu Chan, Shyh-Shin Chiou, Shih-Chang Chuang, Shang-Jyh Hwang

#### Resources

Chien-Hung Chen, Ya-Chung Tian, Chia-Ling Chen, Yao-Fan Fang, Ji-Tseng Fang, Yi-Hao Yen, Wei-Chi Wu, Wen-Shih Huang, Chi-Chin Sun, Wen-Chien Chou, Ching-Hung Lin, Tsung-Hua Yang, Pei-Lin Lee, Ming-Yang Wang, Tsen-Fang Tsai, Tung-Hung Su, Jyh-Ming Liou, Shun-Fa Yang, Chia-Chuan Hsieh, Chih-Chien Sung, Feng-Chih Kuo, Shih-Hua Lin, Dueng-Yuan Hueng, Chien-Jung Lin, Hueng-Yuan Shen, Chang-Hsun Hsieh, Shinn-Zong Ling, Tso-Fu Wang, Tsung-Jung Ho, Pei-Wei Shueng, Chen-Hsi Hsieh, Kuo-Shyang Jeng, Gwo-Chin Ma, Ting-Yu Chang, Han-Sun Chiang, Yi-Tien Lin, Kuo-Jang Kao, Chen-Fang Hung, I-Mo Fang, Po-Yueh Chen, Kochung Tsui

#### Data Curation

Ming-Fang Tsai, Erh-Chan Yeh, Yi-Jung Lin, Yu-Chuang Huang, Wan-Ru Li

#### Writing - Original Draft

Pui-Yan Kwok, Yen-Tsung Huang, En-Yu Lai, Jia-Ying Su

#### Writing - Review & Editing

Jer-Yuarn Wu, Chun-yu Wei, Ling-Hui Li, Cathy S.-J. Fann, Hsin-Chou Yang, Chien-Hsiun Chen, Hung-Hsin Chen, Yi-Min Liu, Ming-Fang Tsai, Erh-Chan Yeh

#### Supervision

Pui-Yan Kwok, Jer-Yuarn Wu, Wei-Jen Yao, Shiou-Sheng Chen,Ming Chen, Chih-Yang Huang, Da-Wei Wang, Chun-houh Chen

#### Project administration

Yi-Min Liu, Tsai-Chuan Chen, Wei-Ting Huang,

#### Funding acquisition

Pui-Yan Kwok

