## Supplement for "Deleterious Variants Contribute Minimal Excess Risk in Large-Scale Testing"

A separate attachment named "Supplementary Table 1.csv"

Table S1: **Selected 292 deleterious variants.** We listed dbSNP ID, Affymetrix ID, chip information, variant location, gene name, inheritance patterns, minor allele frequency, affected numbers in each allele type, associated disease, phenotype category, prevalence, penetrance, risk difference, risk ratio, $p$-value, false discovery rate, and annotated databases.

| | | TPM1 | | TPM2 | | Union | |
|---|---|---|---|---|---|---|---|
| Total | | 142,480 | | 306,968 | | 444,465 | |
| Sex | Female | 78,243 | (54.9%) | 170,768 | (56%) | 246,181 | (55%) |
| | Male | 64,237 | (45.1%) | 136,200 | (44%) | 198,284 | (45%) |
| Age | 0-20 | 1,440 | (1.0%) | 4,685 | (1.5%) | 6,025 | (1.36%) |
| | 20-30 | 8,971 | (6.3%) | 24,063 | (7.8%) | 32,606 | (7.34%) |
| | 30-40 | 16,514 | (11.6%) | 36,287 | (11.8%) | 52,197 | (11.74%) |
| | 40-50 | 21,826 | (15.3%) | 51,496 | (16.8%) | 72,500 | (16.31%) |
| | 50-60 | 25,671 | (18.0%) | 58,733 | (19.1%) | 83,588 | (18.81%) |
| | 60-70 | 32,462 | (22.8%) | 67,361 | (21.9%) | 98,722 | (22.21%) |
| | 70-80 | 23,667 | (16.6%) | 45,517 | (14.8%) | 68,413 | (15.39%) |
| | ≥80 | 11,929 | (8.4%) | 18,826 | (6.1%) | 30,414 | (6.84%) |

Table S2: **Demographic characteristics of the TPMI Biobanks.** The numbers reflect individuals in each respective cohort after quality control (QC) procedures were applied. The column labeled “Both” represents the combined and QC-ed results of individuals from both TPM1 and TPM2 cohorts. For individuals present in both cohorts, genotyping results from TPM2 were preferentially used. The table details the distribution by sex and age group, indicating both absolute numbers and percentages of the total cohort.

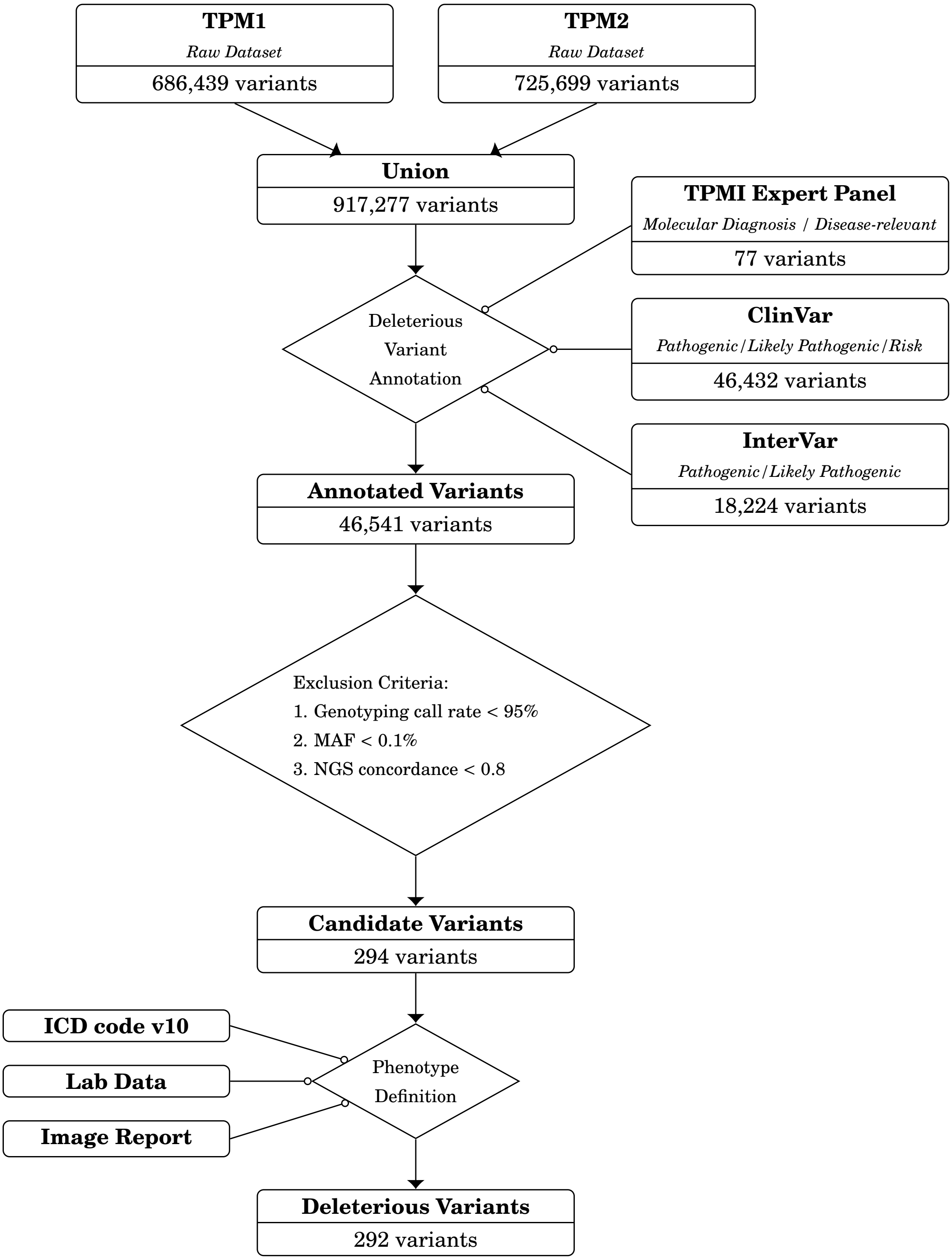


Figure S1: **Variant curation of the 292 deleterious variants.** The candidates of actionable variants were recruited from Clinvar, InterVar, and the common disease variants suggested by TPMI expert panel. The filter criteria were MAF ≥ 0.1%, next generation sequencing (NGS) concordance ≥ 80% in either TPM1 or TPM2 dataset. The diseases without accurate ICD codes were also removed. The concordance rate between SNP genotyping and NGS data was defined by balanced accuracy, i.e., (sensitivity + specificity)/2.

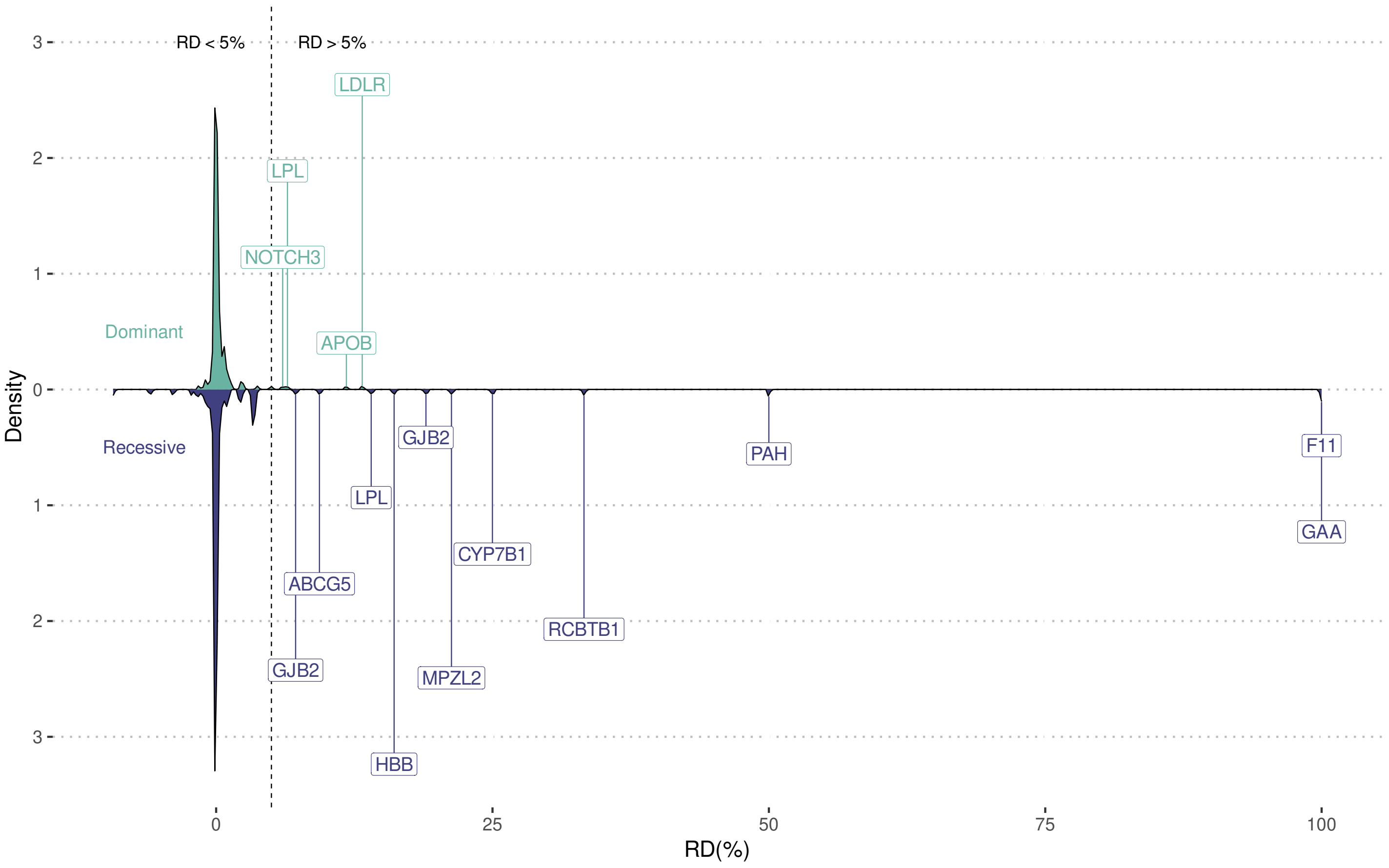


Figure S2: **RD Distribution of the 292 deleterious variants.** We showed the RD distribution of dominant and recessive patterns. We labeled the gene names on the 15 variants with > 5% RD.

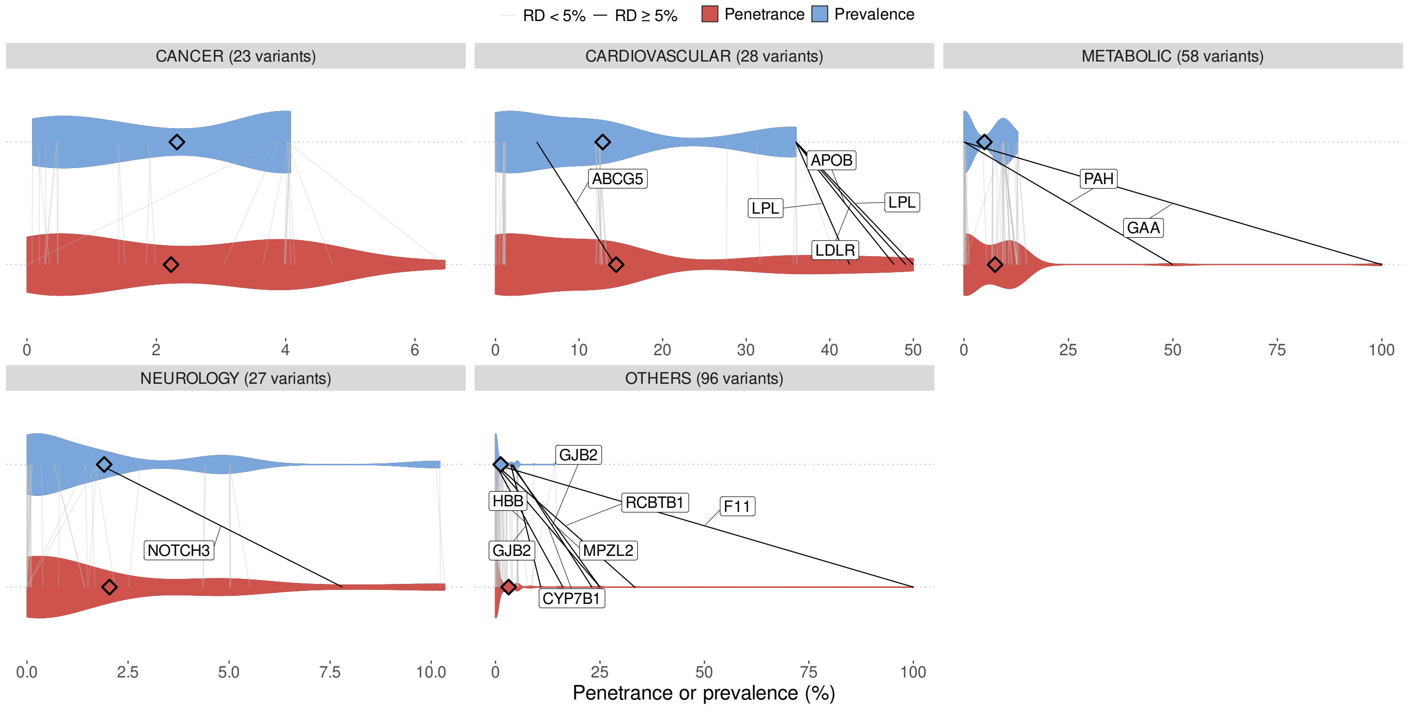


Figure S3: **Violin plots of the 292 pathogenic variants.** To better understand the trends exhibited by each genetic variant across different classifications, we generated violin plots for various phenotype categories. These plots are enhanced with diamond symbols to denote the means within each category and color-coded for clearer differentiation: variants with an RD $\geq 5\%$ are marked with black solid lines and labeled with the corresponding gene names, while those with an RD $< 5\%$ are indicated with light gray solid lines. The width of each violin plot reflects the density of observations in that region, providing a visual representation of data distribution. All violin plots were standardized to a uniform maximum width.

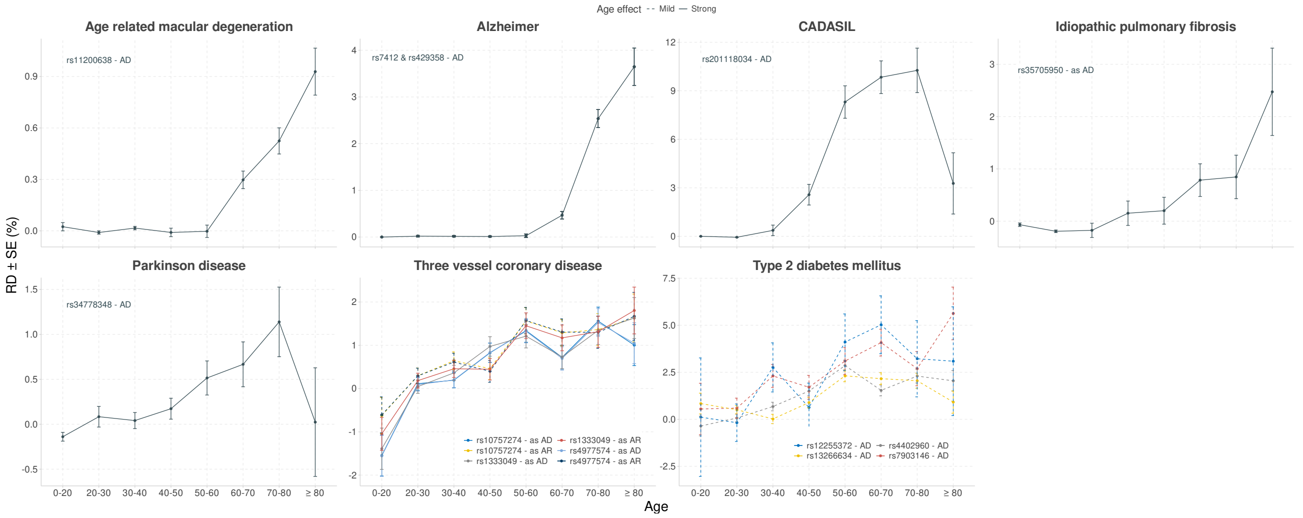


(a) Increasing

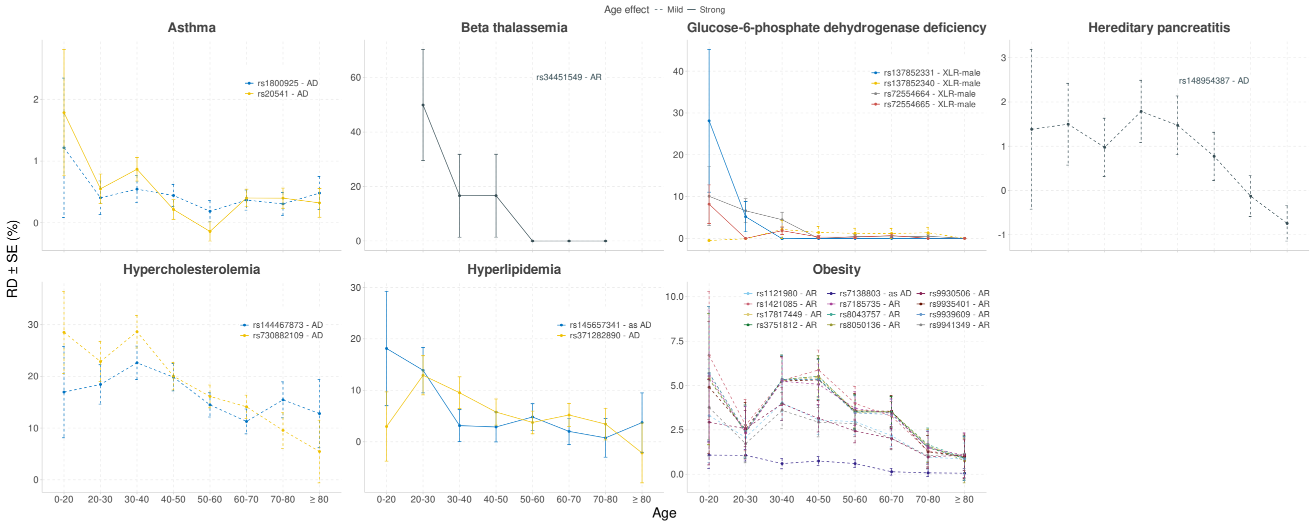


(b) Decreasing

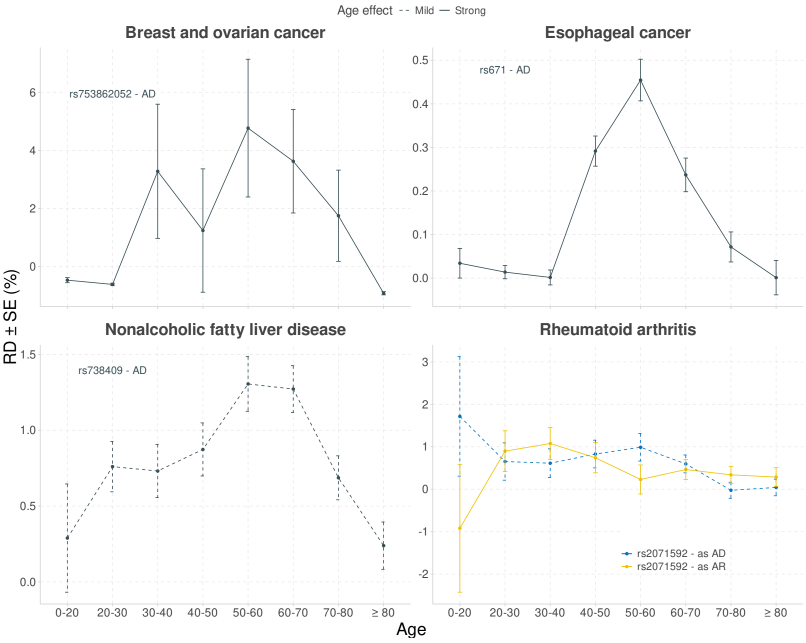


(c) Bell-shaped

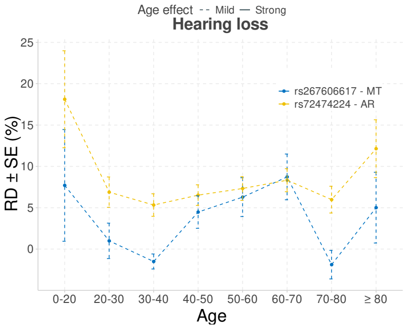


(d) Flat

Figure S4: **Age-stratified RD patterns.** We exhibited the variance in RD as a function of age to visualize the relationship between them. We first selected variants with RD $\geq 5\%$ or FDR $< 0.01$, then filtered out those with less than 30 carriers to ensure the sample size among age groups. Here, (a) shows diseases with increasing RDs with age, indicating rising susceptibility; (b) depicts diseases with decreasing RDs, suggesting higher risks at younger ages; (c) presents a bell-shaped distribution, peaking at mid-life; (d) displays diseases with consistent RDs across all ages, implying uniform genetic impact. Each variant was analyzed for RD scale, defined as $|\max(\mathrm{RD}) - \min(\mathrm{RD})|/\mathrm{median}(\mathrm{RD})$. Diseases with an RD scale $\geq 3$ are depicted with solid lines to indicate a strong age effect, emphasizing substantial fluctuations in risk. Conversely, diseases with an RD scale $< 3$ are depicted with dashed lines, indicating a mild age effect.

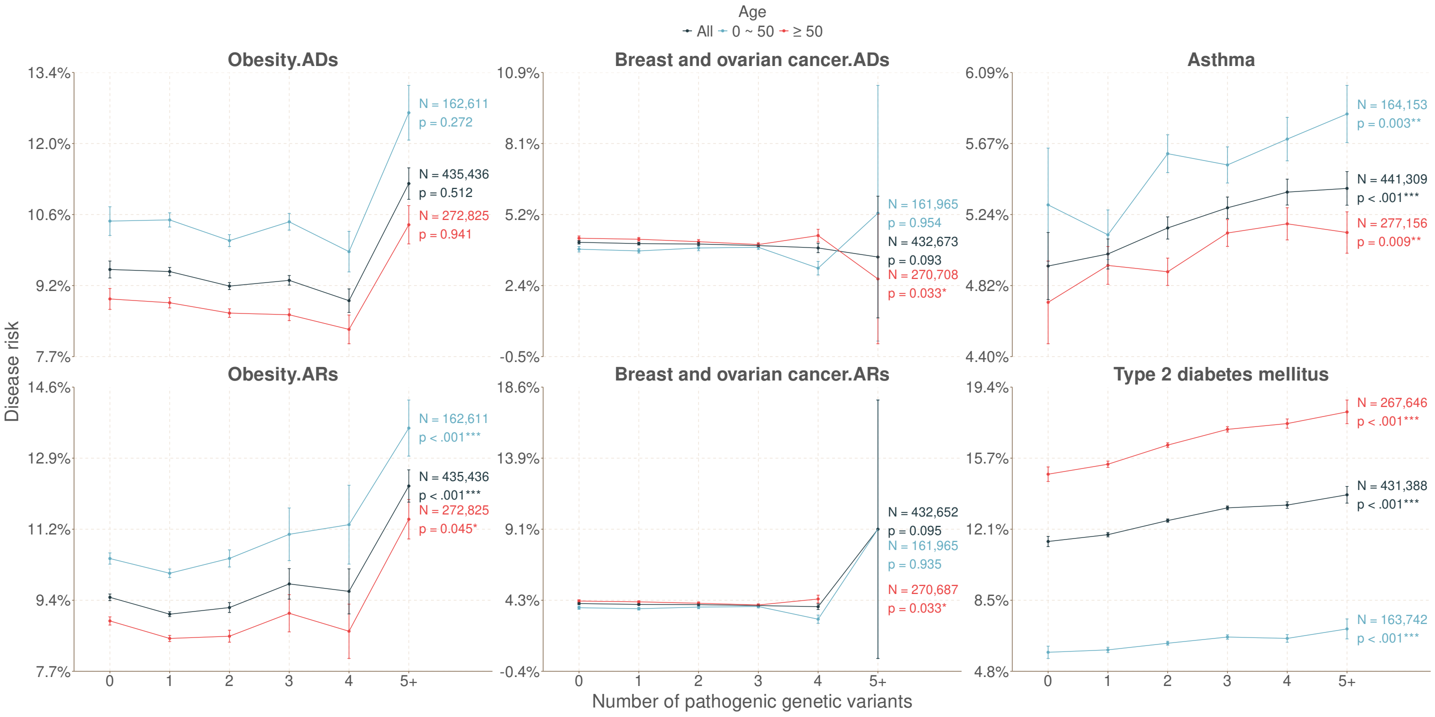


Figure S5: **Dose-response relationship.** We performed dose-response analyses for diseases associated with more than five genetic variants, resulting in the selection of six specific diseases: obesity, breast and ovarian cancer, asthma, type 2 diabetes mellitus, glucose-6-phosphate dehydrogenase deficiency, and retinitis pigmentosa. The Cochran-Armitage trend test was utilized to ascertain whether there is a significant relationship between variant counts and increased disease risks. In the analyses of phenotypes such as obesity and breast and ovarian cancer, some variants display unclear inheritance patterns. Consequently, these variants were analyzed under both autosomal dominant (AD) and autosomal recessive (AR) scenarios, resulting in the distinctions between ".ADs" and ".ARs" in the figure. Diseases such as glucose-6-phosphate dehydrogenase deficiency and retinitis pigmentosa were excluded from this analysis due to insufficient patient data to ensure reliable statistical interpretation.
